# Which Components of the Haemodynamic Response to Active Stand Predict Cardiovascular Disease and Mortality? Data From The Irish Longitudinal Study on Ageing

**DOI:** 10.1101/2024.07.30.24311251

**Authors:** Belinda Hernández, Adam H. Dyer, Cathal McCrory, Louise Newman, Ciaran Finucane, Rose Anne Kenny

## Abstract

**Background:** An integrated haemodynamic response during standing may serve as an integrative marker of neuro-cardiovascular function. Individual components of both heart rate (HR) and blood pressure (BP) responses to active stand (AS) have been linked with cardiovascular disease (CVD) and mortality. We assessed longitudinal associations between entire HR/BP response curves during AS, incident CVD and mortality over 12 years.

**Methods:** Beat-to-beat measurements of dynamic HR/BP responses to AS were conducted in 4,336 individuals (61.5±8.2 years; 53.7% female). Functional Principal Components Analysis was applied to HR/BP response curves and their association with CVD and mortality assessed. We hypothesised that integrating BP/HR information from the entire haemodynamic response curve may uncover novel associations with both CVD and mortality.

**Results:** Higher systolic BP (SBP) before AS and blunted recovery of SBP during AS was associated with all-cause mortality over 12-years (Hazard Ratio [HR]: 1.14; 1.04, 1.26; p=0.007). Higher baseline/peak HR and lower HR from 30 seconds post stand onwards were associated with lower mortality due to circulatory causes (HR: 0.78; 0.64, 0.95; p = 0.013). Higher HR throughout AS was associated with mortality from other causes (HR: 1.48; 1.22, 1.80; p<0.001). Findings persisted on robust covariate adjustment.

**Conclusions:** We observed distinct relationships between HR/BP responses to AS and 12-year incident CVD and mortality. Integrating the entire haemodynamic response may reveal more nuanced relationships between HR/BP responses to AS, CVD and mortality - serving as an integrative marker of neuro-cardiovascular health in midlife and beyond.

## Introduction

Adults typically stand from sitting or lying supine between 50-60 times per day (1, 2). The haemodynamic response to standing (or orthostasis) is a mild physiological stressor requiring integrated neuro-cardiovascular function, which can be readily assessed clinically through measurement of blood pressure (BP) and heart rate (HR) responses to formal active stand (AS) in the clinic (3, 4). In addition to its role in assessing for orthostatic hypotension (OH) and autonomic dysfunction in older adults, the haemodynamic response to active stand can give an excellent indicator of the integrity of short-term cardiovascular and neurogenic responses (5–7).

OH is defined as a drop of ≥20 mmHg systolic/ ≥10 mmHg diastolic BP on standing from supine. In addition to being a significant cause of falls and syncope in older adults (8–10), OH has been associated with cardiovascular/cerebrovascular disease (11–17), cognitive impairment/dementia (18–21), depression (22), gait impairment (23) and frailty (24, 25). These associations highlight the role of haemodynamic response to active stand as an important indicator of intact homeostatic neuro-cardiovascular function and as a potentially important marker of physiological health in older adults.

Importantly, OH and impaired BP responses to AS have also been associated with an increased risk of mortality in community-dwelling older adults (11, 26–32), although negative studies also exist (33, 34). Nearly-all studies have used sphygmomanometer measurement of BP at discrete timepoints, such as 40 or 60 seconds post-stand. Such measurements fail to account for the broader range of haemodynamic responses to active stand. Broader responses can be readily assessed using non-invasive beat-to-beat finometry, which gives waveform measurements similar to those from invasive intra-arterial monitoring, capturing dynamic changes often missed with other techniques (35–38). Using beat-to-beat finometry also enables classification of different BP phenotypes in response to AS which may differ in their associations with important clinical outcomes such as mortality (38, 39).

In addition to BP response to AS, dynamic changes in HR response to AS may also be associated with mortality. Research has clearly demonstrated an association between HR Recovery (HRR) following exercise and increased mortality (40). However, studies vary in stressor applied (most frequently treadmill exercise) and typically assess recovery at 1-5 minute period post-exercise cessation (41, 42). Given that standing is a potent and ubiquitous stressor which can be performed by anyone who is functionally mobile, comparatively few studies have examined the association between HR responses to AS and subsequent mortality in community-dwelling older adults (43, 44).

HR rapidly increases in the immediate period post-stand, peaking at around 10 seconds in order to counteract gravitational forces on BP. This is felt to result from an abrupt inhibition of vagal activity, after which HR rapidly declines reflecting parasympathetic reactivation (45, 46). HR peak and decline typically takes place in the immediate 30-second period post-stand. Previous work has demonstrated that a slower speed of HRR may be a strong predictor of mortality in older adults (47). Similarly, studies have demonstrated significant associations between higher baseline HR before AS and in the 30-60 second post-stand time period and mortality (48). Whilst the association between an increased resting HR and both overall and cardiovascular mortality has been firmly established in the literature (49–51), the relationship of HRR to AS and mortality has been less well explored.

Despite associations between both BP and HR responses to AS and increased mortality in older adults, no study to date has integrated these responses in the same study. Whilst most studies consider BP/HR responses at a single timepoint/scalar summaries of responses at short intervals, none have considered the complete haemodynamic response curve to AS. In the current study embedded within The Irish Longitudinal Study of Ageing (TILDA), we used functional Principal Components Analysis (FPCA) to examine the determinants of variability in continuous beat-to-beat haemodynamic response to AS and their association with both 12-year incident CVD and 12-year mortality. We also show the flexibility and utility of our FPCA model to predict CVD on a separate clinical cohort validation study of patients from the Falls and Syncope Unit from St James’ Hospital, Dublin.

## Methods

### Study Approval, Setting and Participants

#### The Irish Longitudinal Study on Ageing

Data were obtained from Wave 1 (2010) of The Irish Longitudinal Study on Ageing (TILDA), a prospective nationally-representative study of community-dwelling older adults in Ireland. Participants with dementia and/or cognitive impairment were excluded at baseline. All data from the active stand test were obtained in a health centre with assessments administered by trained nurses (52, 53). The study was approved by the Faculty of Health Sciences Research Ethics Committee at Trinity College Dublin and adhered to the Declaration of Helsinki. A total of 4,336 participants were included in the analysis from an initial sample of 4,956. A flow chart including inclusion criteria are shown in Figure S1.

#### The Active Stand

The Active Stand (AS) test in TILDA was performed by participants who completed the wave 1 TILDA health centre assessment. Briefly, participants lay supine for ∼10 minutes before being asked to stand. They then remained standing for two minutes. The test was supervised by a research nurse who assisted with standing as necessary. Continuous beat-to-beat measurements of HR, systolic blood pressure (SBP), diastolic blood pressure (DBP) and stroke volume were captured at 1Hz from 30 seconds prior to standing to 2 minutes post standing (resulting in 151 measurements per person) using a Finometer MIDI device (Finapres Medical Systems BV, Amsterdam, the Netherlands). A finger cuff was placed on the middle or proximal phalanx of a finger (as per manufacturer instructions) on the left hand. Calibration was performed using Physiocal™, while hydrostatic pressures differences were corrected for using the system’s height correction unit.

Absolute HR was analysed as this is the only signal which can be accurately captured by the Finometer in absolute terms (54), for all other signals change from baseline (60 to 30 seconds before standing) was analysed. SV was normalised by body surface area prior to analysis. Therefore, this signal describes the change from baseline SV per square meter of body surface area and is referred to as the stroke volume index (SVI) henceforth.

#### Mortality Status

In Ireland, all deaths are registered through the General Register Office (GRO). Mortality status and cause of death were determined by inspection of death certificates over a ∼12-year follow-up using data linkage between TILDA and the GRO. The methodology and procedure for determining cause of death has been documented elsewhere (55). Data are available for all reported deaths of TILDA participants up to 31st January 2022. Deaths unofficially reported to TILDA by family members but not yet included in the GRO were excluded as the exact date and cause of death was not ascertainable as were deaths by suicide or deaths due to accidents (n = 29 see Figure 1).

**Figure 1:**
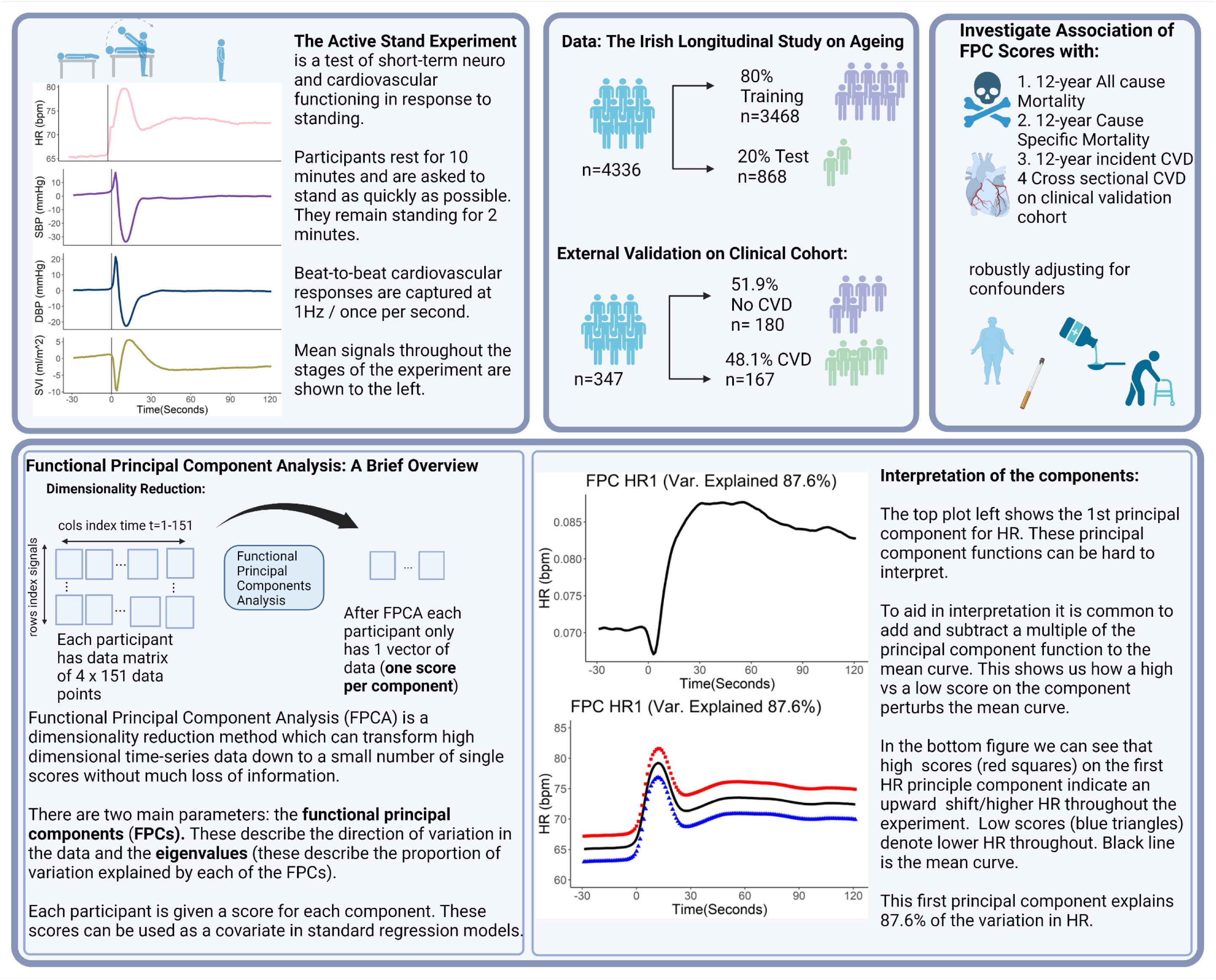
Overview of the methodology used to investigate the association between haemodynamic responses to standing and 12-year mortality and incident cardiovascular disease.

#### Clinical Validation Cohort for Cardiovascular Disease Associations

A cross-sectional clinical validation study of patients referred to the Falls and Syncope Unit (FASU) in St James’ Hospital Ireland was used to assess the association of the functional principal component model for with CVD on an independent cohort. As the study was cross-sectional, mortality data was not available in the replication cohort. Briefly, patients aged 50 and over who had been referred to the FASU due to reporting issues with episodes of dizziness, falls or syncope were recruited into the study. Patients with a diagnosis of cognitive impairment or who were unable to provide informed consent or patients who were unable to perform the active stand experiment were excluded from participation. The study was approved from the Tallaght University Hospital/St. James’s Hospital Joint Research Ethics Committee (REC:2018-08,CA,16).

### Statistical Analysis

#### Univariate Analysis

For univariate descriptive summaries categorical variables were expressed as percentages and compared between groups using a Chi-Squared test. Continuous variables were expressed as a mean and standard deviation and compared between groups using a one-way ANOVA.

For univariate summaries of the peripheral haemodynamic signals, group means were compared using functional ANOVA. For more details on this methodology see Supplementary Material S1.

#### Functional Principal Components Analysis

FPCA is an extension of the well-known dimensionality reduction technique principal components analysis (PCA) for functional data (in this case haemodynamic responses as a function of time). In the following analyses functional principal components analysis (FPCA) was performed separately on each of the peripheral haemodynamic signals. The number of eigenfunctions (principal components) which explained 90% of the variability in the data were retained. A total of 2 functional principal components (FPCs) were retained for HR, 5 for SBP, 6 for DBP and 4 for SVI. FPC scores associated with these components were then extracted and used as covariates in the models for CVD and mortality described in the following section. A brief explanation of FPCA and study design can be seen in Figure 1 see also Supplementary 2 for a more detailed methodological description and (59–61) for further examples.

#### Multivariable Survival Models

A Cox PH model was implemented to model all-cause mortality with age of death as the outcome variable. Those who survived were right censored to their age as of 31^st^ Jan 2022. The survival status of all participants is known as of 31^st^ January 2022 regardless of attrition from the TILDA study due to data linkage with the GRO. Because of the high correlation between response of SBP and DBP to standing, only the results for SBP are shown. However, including DBP in the models does not change the conclusions of this work. The proportionality assumption of the Hazard functions was tested by assessing the scaled Schoenfeld residuals over time.

For cause-specific mortality, a competing risk analysis using the Fine and Grey method was used to model cause specific mortality.

In all cases models were trained on a random 80% of the data and tested on the remaining 20% which was independent of the model training. To assess the model performance of both the all-cause and cause specific models on independent data; the time-dependent AUC and 95% confidence interval was calculated using the 20% test dataset.

#### Elastic Net Penalised Regression and analysis of CVD

CVD in the TILDA study was defined as presence of at least one of the following conditions: congestive heart failure, angina, heart attack or atrial fibrillation at follow up waves 2-6 (∼ 12 year follow up). Participants with CVD at baseline were removed from the analysis. In total 2,419 participants who didn’t have CVD at wave 1 and attended all six TILDA waves and were included in the analysis of incident CVD.

High class imbalance was present for incident CVD (9.6% of participants had incident CVD). For this reason, the model was trained across 1,000 balanced bootstrap samples to ensure that the associations discovered were not due to bias in selecting the control cases. Over each of the 1,000 permutated datasets we employed an elastic net penalised regression which is a common method for variable selection. The optimal λ was chosen using 10-fold cross validation on the training data and α = 0.5. The variables which had non-zero coefficients in more than 50% of the permutations were considered consistently associated with CVD and presented in the results. Again, all performance metrics were calculated based on the 20% test set which was not used in any part of the model building.

In the FASU clinical validation cohort, CVD was defined as the presence of any of the following conditions: transient ischemic attack, stroke, carotid stenosis, ischemic heart disease, angina, myocardial infarction, atrial fibrillation, heart failure, diabetes. To ensure uniformity of methodologies across datasets the same methodology as described for incident CVD above was also used on this cohort. Age, sex, BMI, smoking history, hypertension and high cholesterol were controlled for.

All statistical analysis was performed using the R version 4.2.3.

#### Covariates

The following covariates known to be associated with mortality and incident CVD were controlled for in all models based on TILDA data: sex (binary); education (Primary/Secondary/Third level); smoking history (Never, Former, Current); BMI (numeric); hypertension (binary); high cholesterol (binary), depression, anxiety or psychosis (binary) and frailty.

Hypertension was defined as systolic blood pressure >140 mmHg and/or diastolic blood pressure >90 mmHg and/or use of any of the following medications: ATC codes C02, C03, C07, C08, C09. Presence of high cholesterol was defined as Total Cholesterol ≥5 mmol/L and/or use of ATC C10 (excluding C10AX06). Presence of depression, anxiety or psychosis was defined as use of any of the following medications: N05BA, N05AE, N05CD, N05CF, N06A, N05A) and/or a score ≥11 on the Hospital Anxiety and Depression Scale (HADS-A) and / or a score of ≥16 on the Centre for Epidemiological Studies Depression Scale (CES-D). Frailty was defined using the 32-item frailty index (56) which captures information regarding disability, symptoms, signs and diseases affecting a range of organ systems. The FI has previously been operationalised in TILDA and characterises participants into robust (FI score <0.1), pre-frail (0.1≤ FI score <0.25) or frail (FI score ≥ 0.25) based on their index scores (57, 58).

For all-cause mortality further adjustment for presence of CVD and neurodegenerative disease at baseline were accounted for. Specifically, a history of angina, heart attacks, heart failure, stroke, TIA, diabetes and Parkinson’s were additionally controlled for. These variables were not controlled for in models of cause specific mortality due to high correlation of these conditions with deaths due to CVD. When modelling incident CVD, those with CVD at baseline were removed from analysis.

## Results

### Participants

Overall, 4,366 TILDA participants (mean age 61.5, 53.7% female) were included. During the follow-up time period, 9.7% (n = 422) of the 4,336 eligible participants were confirmed as deceased as of the 31^st^ January 2022. Of these; 116 were due to deaths of the circulatory system, 198 were due to cancer and 108 other causes. Due to sample size limitations deaths due to respiratory illnesses were combined with other causes and so of the 108 mortalities classified as “other”, 31 are known to be due to respiratory illnesses.

### Univariate Summary

Table 1 shows a summary of each of the covariates by mortality status. As can be seen, age, sex female, lower education, hypertension and depression/anxiety/psychosis were all positively associated with all-cause mortality. Only age and hypertension were found to differ according to mortality cause.

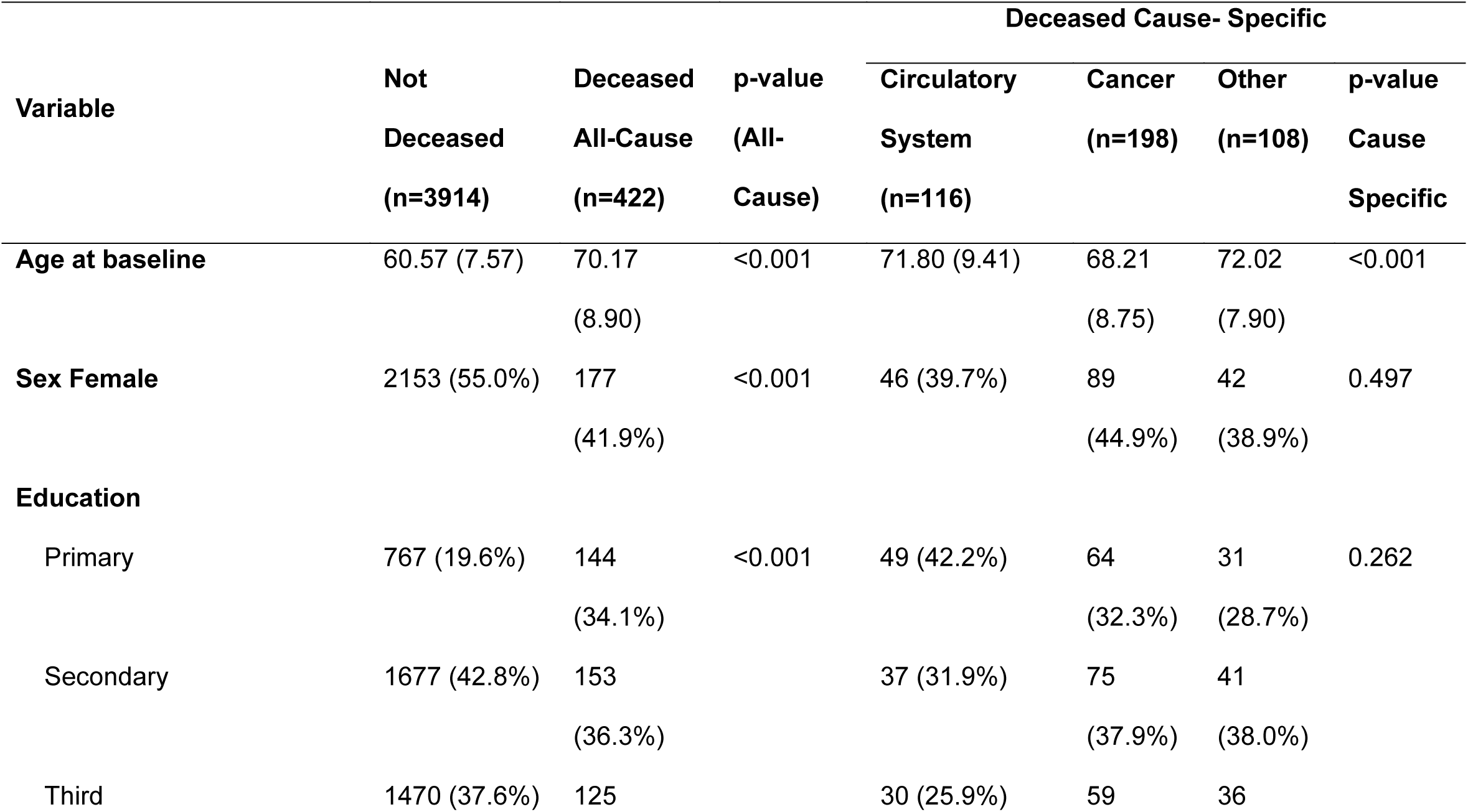
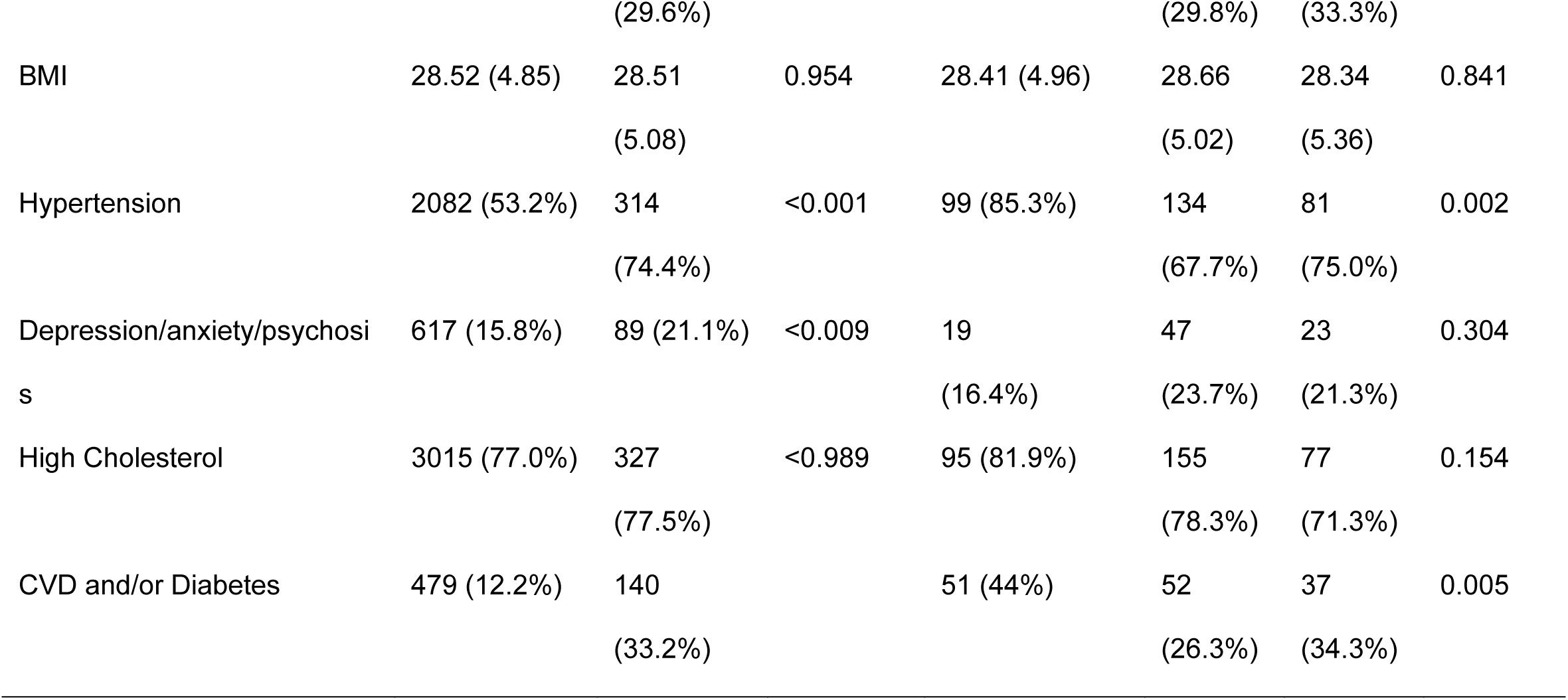

Figure 2 A-D shows the mean trace of each of the haemodynamic signals according to survival status for all-cause mortality. As can be seen, there is evidence to suggest that the mean trace of HR, SBP, DBP and SVI differ according to survival status. Specifically, the mortality group have HR that is characterised by elevated supine baseline, a blunted peak on standing and a more gradual recovery between 10-40 seconds post standing (Figure 2A). Recovery of both SBP and DBP in this group is muted between the post-stand nadir and ∼70 seconds post standing (Figure 2 B, C). Change from baseline SVI is characterised by a higher post stand peak and elevated post-standing values in the recovery period from 30 seconds onwards in the mortality group (Figure 2 D).

**Figure 2.**
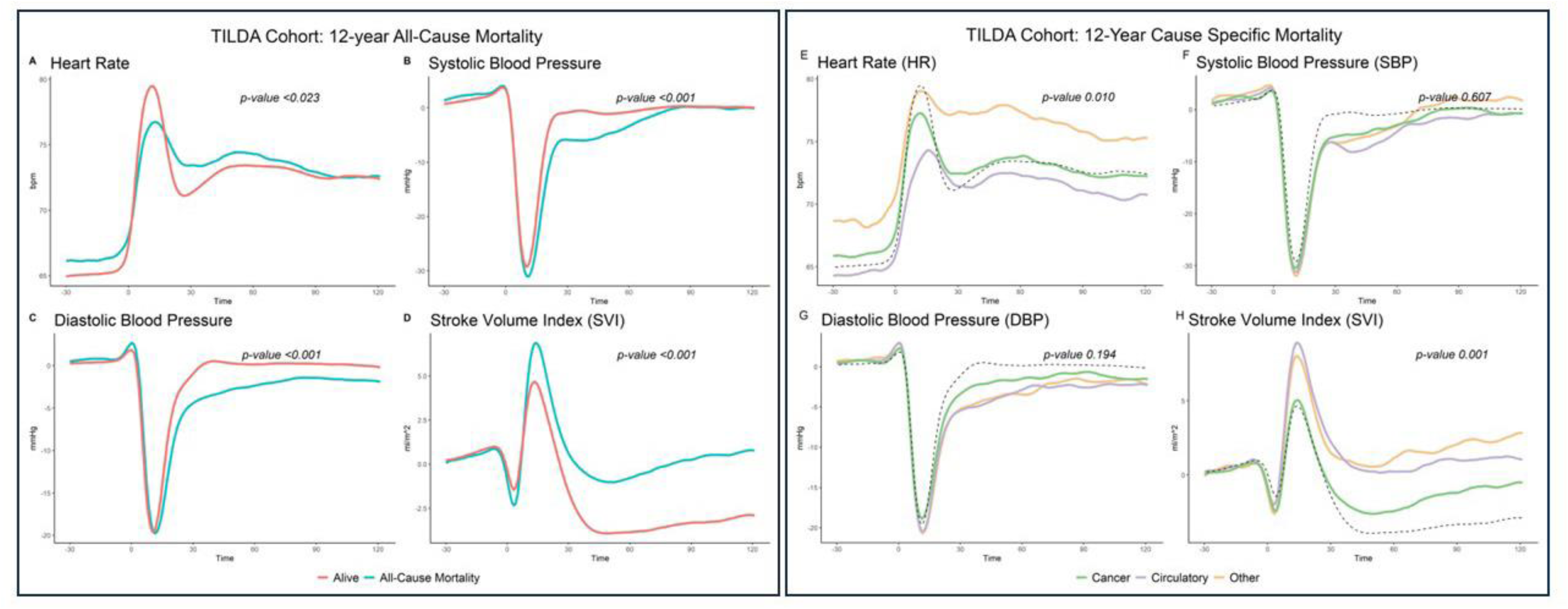
Left Panel (A-D) Mean trace of haemodynamic signals according to All-Cause Mortality survival status. Time 0 denotes stand time. Right Panel (E-H) Mean trace of haemodynamic signals according to Cause-Specific Mortality status. Dashed black line indicates the mean curve for participants who were alive at the last day of follow up 31^st^ January 2022 for reference. Time 0 denotes stand time.

The p-value in each case is the result of a functional ANOVA analysis using a permutation test on a basis function representation of the data. As a sensitivity analysis, several types of ANOVA tests were conducted (with varying assumptions regarding the distribution of the data and structure of the covariance matrix) and found not to change the conclusions of the analysis (see Tables S1 and S2 for full results).

Figures 2 (E-H) depict the mean trace of each of the haemodynamic signals according to mortality cause. As can be seen, only HR and SVI differ significantly between the three recorded causes of mortality (p-value 0.01 and 0.001 respectively). Deaths of the circulatory system have a blunted HR peak and recovery to standing (Figure 2E) as well as an elevated change from baseline SVI from post standing peak onwards compared to deaths due to cancer (Figure 2H). The haemodynamic response for cancer mortality was in general more similar to those who survived 12-year follow-up when compared to circulatory and other causes (Figures 2 E-H). Other mortality was characterised by elevated supine baseline and post-stand HR, as well as a poorly defined peak HR and recovery (Figure 2E). The SVI response for other mortality was similar to HR.

### Multivariable analysis of 12-year all-cause mortality

Figure 3A shows the Hazard Ratio and 95% confidence intervals for the FPC scores associated with all-cause mortality. Figure 3B depicts the mean HR signal and how high versus low FPC scores for the first HR component (FPC HR1) perturb the mean signal. Figure 3C shows the equivalent information for the fifth SBP component (FPC SBP5). Figures 3 B and C can be used to aid interpretation regarding which features of the FPC’s may be discriminating mortality status. To illustrate, the Hazard ratio for each standard deviation increase in FPC SBP5 is 1.13 (95% CI 1.02-1.24, p=0.016) (Figure 3A). Hence FPC SBP 5 is positively associated with all-cause mortality. High scores on this component (red squares, Figure 3B) appear to discriminate higher baseline SBP and possibly blunted recovery of SBP from ∼20-60 post stand. Similarly the scores related to component FPC HR1 were poitively associated with mortality. As can be seen higher scores on this component are associated with elevated heart rate throughout the experiment (red squares, Figure 3C).

**Figure 3:**
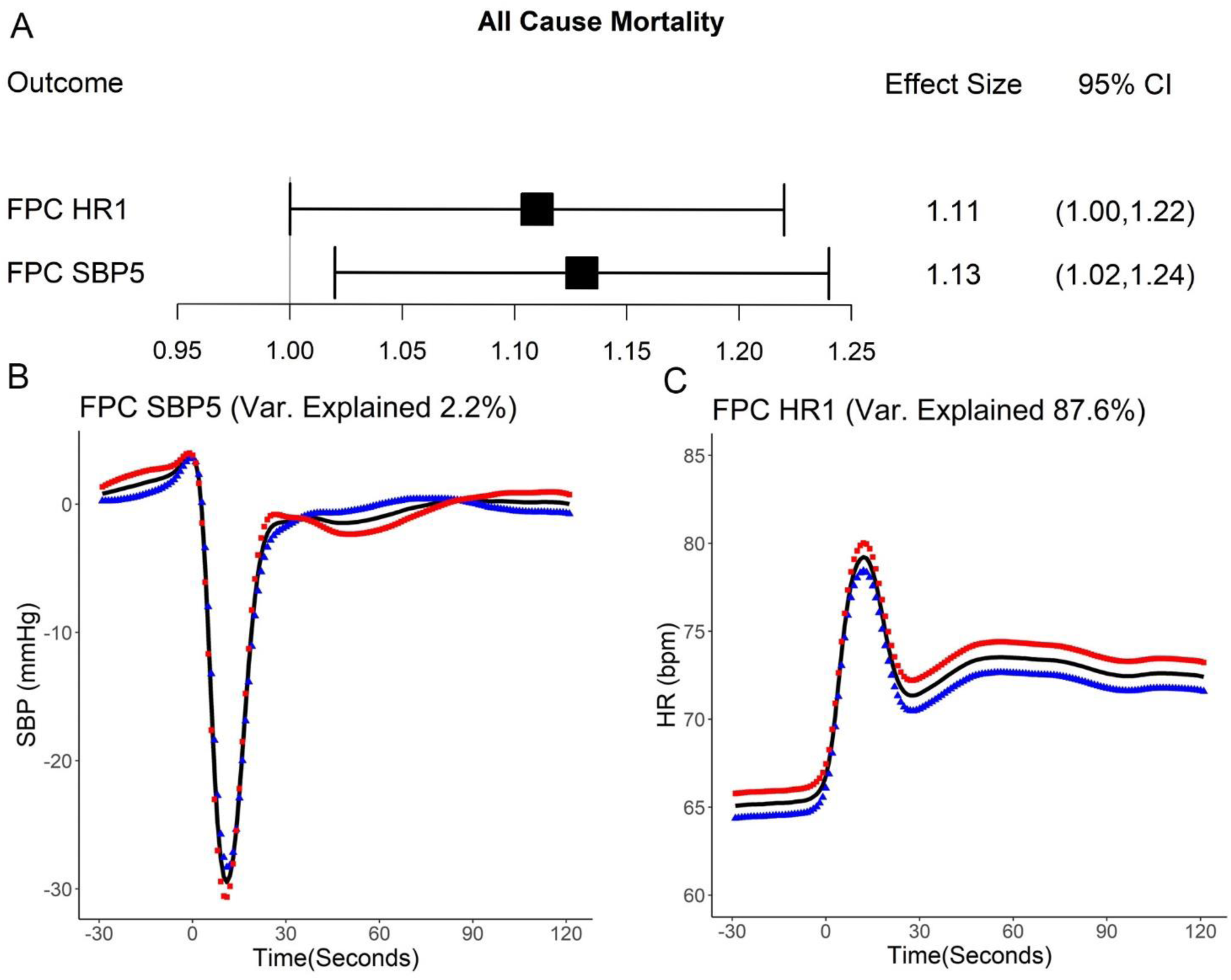
(A) Hazard ratio and 95% CI for the coefficients included in the model for all-cause mortality (B) Modes of variation associated with all-cause mortality (mean signal +/- 30 x eigenfunction (principal component)). (B) Mode of variation for the 5^th^ Systolic Blood Pressure principal component (C) Mode of variation for the 1^th^ Heart Rate principal component. Red squares represent how high scores on the indicated FPC affect the mean, blue triangles represent how low scores on the principal component affect the mean. The mean curve for each signal is indicated by the continuous black line.

### Multivariable analysis for Cause-Specific Mortality

Figures 4 A-D show how the principal components associated with cause-specific mortality affect their respective mean curves. With respect to cause-specific mortality it can be seen from Table 2 that the 2nd principal component for HR was negatively associated with cardiovascular mortality (p-value 0.013) while the 5^th^ component of SBP was marginally positively associated (p-value 0.084). From Figure 4B, lower values on FPC HR2 (blue triangles) discriminate lower baseline and peak HR and higher HR from 30 seconds post stand onwards. Lower heart rate throughout and in particular around the HR peak was indeed a feature of cardiovascular mortality (seen in Figure 2 E).

**Figure 4:**
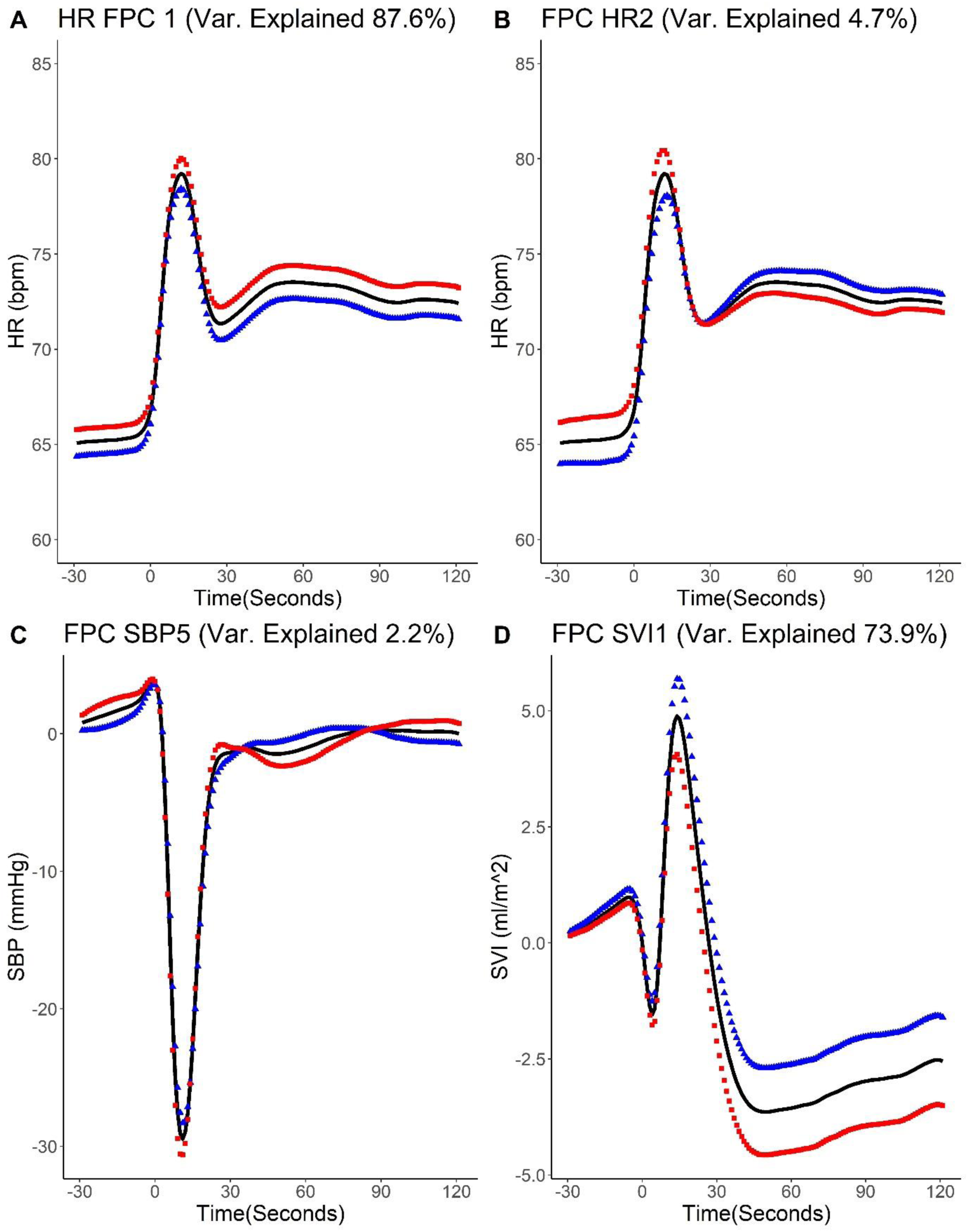
Modes of variation for components associated with cause specific mortality. Red squares represent how high scores on the indicated component affect the mean, blue triangles represent how low scores on the component affect the mean.

**Table 2.** Results of competing risks model for cause specific mortality.

| Model | Hazard Ratio | LCI | UCI | p-value |
| --- | --- | --- | --- | --- |
| **Circulatory System** |  |  |  |  |
| FPC HR2 | 0.78 | 0.64 | 0.95 | 0.013 |
| FPC SBP5 | 1.15 | 0.98 | 1.35 | 0.084 |
| **Cancer** |  |  |  |  |
| FPC HR2 | 1.19 | 1.01 | 1.40 | 0.037 |
| FPC SVI1 | 1.19 | 1.02 | 1.39 | 0.025 |
| **Other** |  |  |  |  |
| FPC HR 1 | 1.48 | 1.22 | 1.80 | <0.001 |
| FPC SBP5 | 1.28 | 1.07 | 1.53 | 0.007 |
| FPC SV1 | 0.68 | 0.55 | 0.85 | <0.001 |

Both FPC HR2 and FPC SVI1 were positively associated with cancer mortality. Respectively these discriminate a higher baseline HR, a higher and pronounced HR peak (red squares Figure 4 B) and lower change from baseline SVI from the post stand maximum onwards (Figure 4 D) which are also features of cancer mortality identified in Figure 2 E and H.

Other deaths were characterised by higher values on FPC HR1 and SBP5 along with lower values on FPCSV1 which is associated with impaired recovery of SPB from ∼30-70 after stand; elevated SVI in the post stand recovery period as well as higher HR throughout the procedure (Figures 4 A,C and D).

### Sensitivity Analysis: Beyond consensus measures

As SBP5 was associated with all-cause as well as cause specific mortality and the major characteristics of SBP5 discriminate high baseline SBP and poor recovery of SBP from 30-70 seconds post stand; we wanted to ensure that our FPC scores were not merely acting as surrogates for baseline SBP and orthostatic hypotension. To investigate this, we performed a sensitivity analysis to examine whether our dynamic measures performed any better than using consensus definitions of orthostatic hypotension and baseline SBP. Hence the above models were repeated replacing the FPC scores with baseline SBP and with binary indicators of orthostatic hypotension (OH) at 10 second intervals from 10 to 60 seconds after standing. Interestingly there was no association between OH and deaths of the circulatory system, deaths from cancer nor other deaths. Regarding all-cause mortality only OH40 was significantly associated. Baseline SBP was not predictive of all-cause nor cause specific mortalities (see Table S5 for full results).

### Model accuracy

The area under the ROC curve (AUC) as a function of age at death for all-cause and cause specific mortality are depicted in Figure 5. In all cases the cross-validated AUC is based on 20% test data which was not used to train the model. The AUC for all-cause mortality was quite stable across age and ranged from 91.2% (age 67) to 75.9% (age 87). With respect to circulatory deaths, the AUC more accurately discriminated mortality at younger ages (from 65-71) than those at older ages. There was good discrimination of cancer mortalities from ages 65-83 was (average AUC 86.47%) with wider confidence intervals beyond that age range while other mortalities could not be accurately discriminated (95% CI overlapped 50% in nearly all cases). The improved predictive accuracy for cancer mortality may perhaps be due to this being the most common cause of death in our sample (46.9% of deaths compared to 27.5% due to deaths of the circulatory system and 25.6% due to other causes).

**Figure 5:**
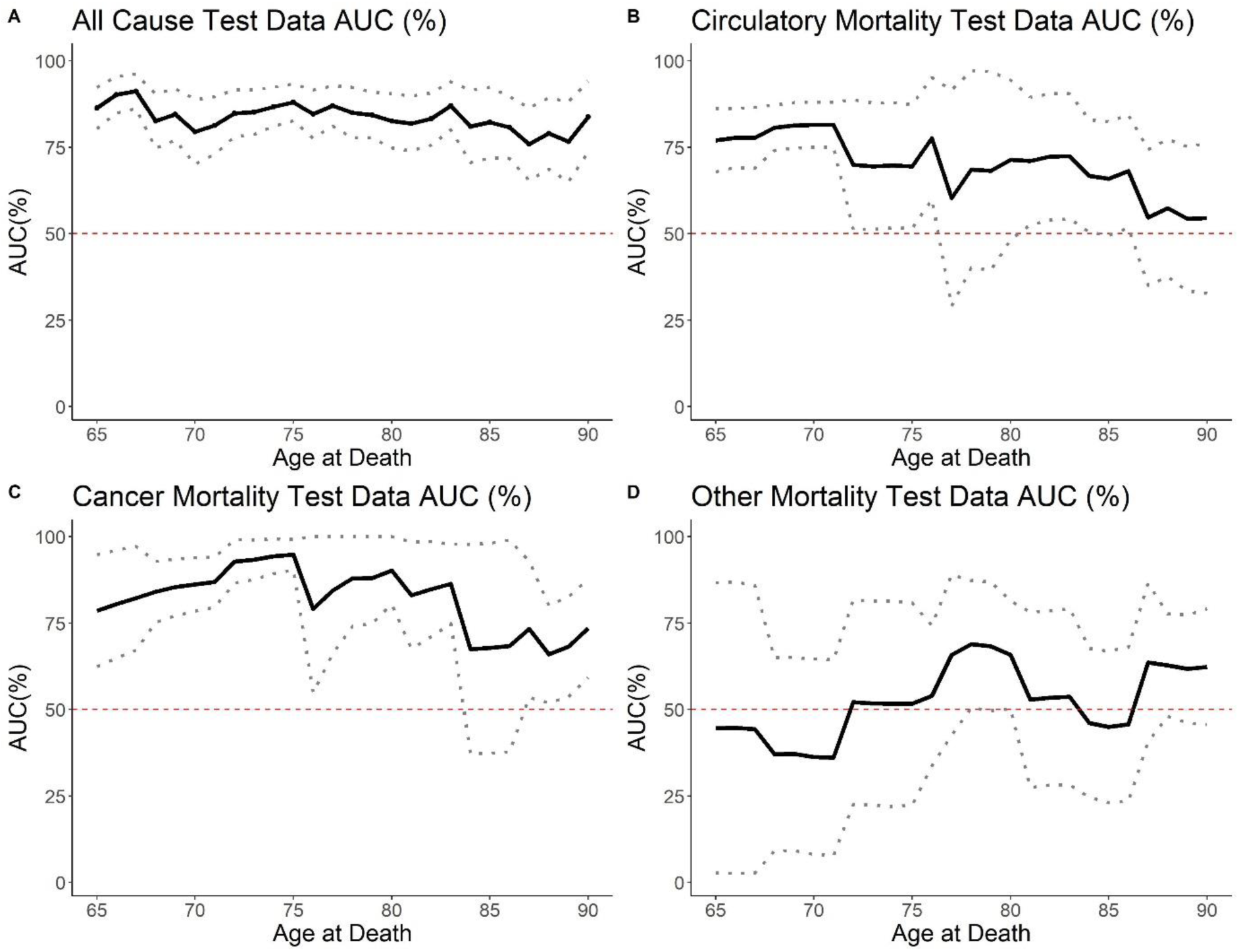
Estimated time-dependent AUC and 95% CI according to age at death on 20% hold-out test data (A) All-Cause Mortality, (B) Circulatory Mortality, (C) Cancer Mortality and (D) Other Mortality.

### Investigating 12-year incident CVD

Figure 6 shows the mean baseline haemodynamic profiles of participants with 12-year incident CVD (n=229) versus those who did not have CVD after 12-year follow up (n=2182). Those with a history of CVD at baseline were removed from the analysis. As can be seen HR, DBP and SV were significantly associated with 12-year incident CVD status in univariate analysis (Figures A,C,D, p-value 0.038,0.001 and <0.001 respectively). Those with incident CVD had on average lower HR throughout the experiment and in particular a blunted recovery to standing around the HR peak, followed by lower HR in the recovery from 30-180 seconds after standing. They also had a significantly lower DBP/blunted recovery of DBP on standing in the recovery period from ∼30 seconds after stand onwards as well as an elevated change from baseline SVI from the initial response to standing ∼5 seconds after standing to the end of the experiment.

**Figure 6:**
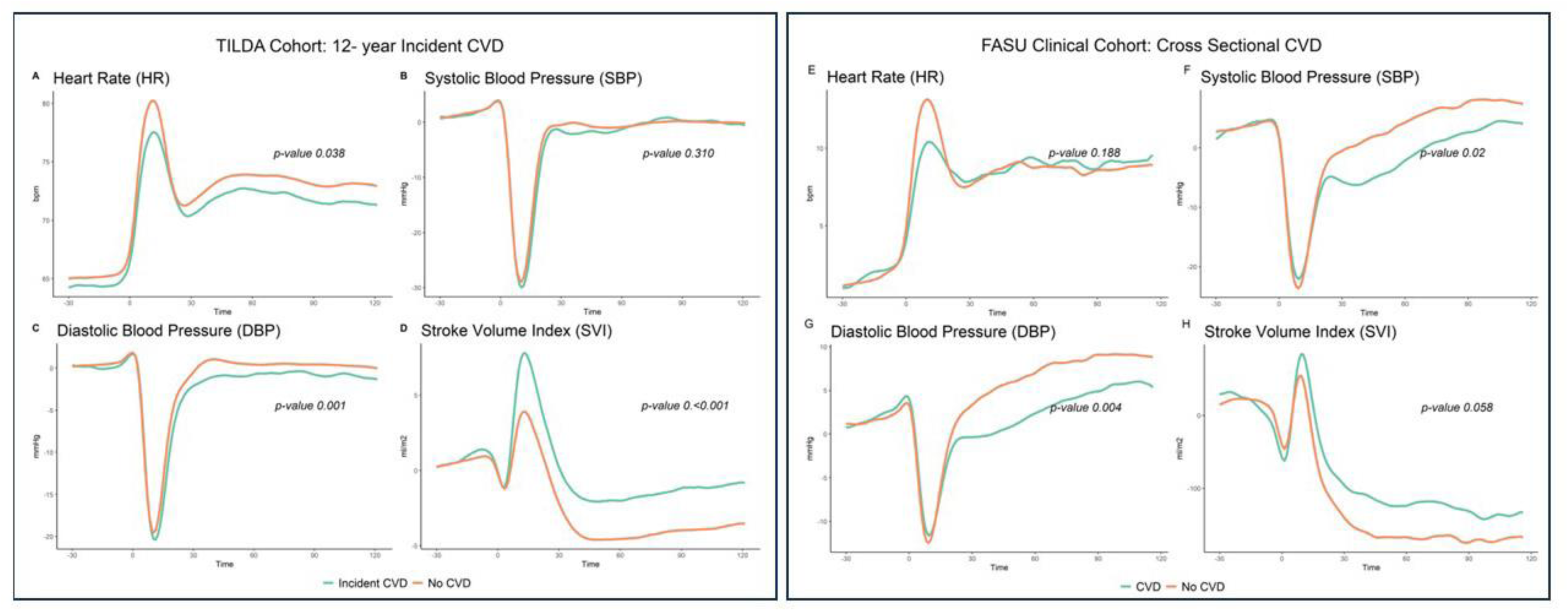
Left Panel A-D: Baseline haemodynamic responses to standing for participants who had no history of CVD (wave1 2009-2010). Groups denote participants who subsequently went on to have incident CVD vs those who did not have CVD during 12-year follow-up. Right Panel E-H Mean haemodynamic responses to standing for patients with diagnosed CVD vs No CVD in a separate clinical cohort.

With regards to multivariable analysis; after adjusting for age, sex, education, medication usage, BMI, smoking and frailty status, the FPC scores for HR1 and SV1 remained significantly associated with incident CVD status. The components associated with incident CVD were related to lower HR and elevated post stand SVI which was in line with the univariate analysis depicted in Figure 7 (see Figure S2 for mean trace and how high versus low scores on the FPCs significantly associated with incident CVD perturb this mean curve. See also Table S6 for the odds ratio and 95% CI of variables significantly associated with incident CVD). Regarding model accuracy, the mean AUC based on 20% test data was 68.9%, 95% CI (62.6%,73.5%); sensitivity 61.2% and specificity 65.6%.

### External Validation on a clinical cohort

The FPC models trained on the TILDA study were exported and validated on a separate clinical cohort (n=347; 56.2% female; age mean (sd) 70.4 (10.3)) from the Falls and Syncope Unit in St James’ Hospital. Of the 376 total patients 167 (48.1%) had CVD. Figure 6 E-H shows the mean haemodynamic curves for those with CVD vs no CVD for this cohort.

After adjustment for age, sex, BMI, smoking history, and medication usage the second FPC for diastolic blood pressure which discriminates a blunted drop in blood pressure to the nadir and impaired blood pressure recovery from 35 seconds onwards remained significantly associated with presence of cardiovascular disease (see Fig S2). The AUC on the 20% test data was 0.64 95% CI (0.56,0.71), sensitivity 0.63, specificity 0.60.

## Discussion

In this study of over 4,300 community dwelling older adults, we have shown that functional principal component scores of haemodynamic responses to standing were independently associated with 12 year all-cause and cause specific mortality and also independently associated with 12-year incident CVD even after robust adjustment for health, frailty, behavioural and demographic risk factors (average cross validated AUC 83.7%,81.8% and 70.0% for all-cause, cancer and circulatory system mortality respectively; AUC 68.9% for incident CVD). We also showed that recovery of DBP was associated with cross sectional CVD in a separate clinical cohort.

Briefly, all-cause mortality was associated with principal components which discriminated elevated baseline SBP and impaired SBP recovery from ∼30-60 seconds after standing as well as higher HR in general. Mortality of the circulatory system was associated with principal components of HR which discriminated lower supine baseline HR and blunted HR peak as well as impaired recovery of SBP ∼30-70 seconds after stand. Cancer mortality was associated with components which discriminated lower change from baseline SVI from ∼10 seconds post stand onwards and a pronounced/well defined HR peak. These characteristics of cancer mortality were more similar to those who survived than to those who died of CVD and other causes. Similar to circulatory system mortality, other mortalities were also associated with components which discriminated impaired SBP recovery as well as elevated HR and SVI in recovery to standing. Meanwhile, incident CVD was associated with lower HR throughout and an elevated SVI in the initial and sustained response to standing from ∼5 seconds post stand to the end of the experiment.

Many of these findings are in line with previous research. In particular, (47) discovered that slower speed of heart rate recovery from 10-20 seconds after standing was associated with all-cause mortality and likely reflected dysregulation of the response of the parasympathetic nervous system; as parasympathetic inhibition in the immediate response to standing and subsequent reactivation in the initial recovery ∼10 seconds after standing are thought to be responsible for the peak and subsequent drop in HR in response to orthostasis. The work of (62) and others also found that higher resting heart rate was positively associated with mortality (see also 49, 50).

Supine hypertension either on its own or with concomitant OH has previously been associated with shorter survival and higher risk of cardiovascular events as it represents a dysfunctional response in the autonomic reflexes required to maintain blood pressure upon standing (63, 64). Other studies have found evidence of U shaped association with respect to supine blood pressure and age where lower supine SBP was associated with mortality in those aged 85+ (65). Interestingly, our results showed that neither OH from 10-60 seconds post stand nor resting supine SBP were significantly associated with 12-year all cause nor cause specific mortality (the only exception being OH40 for all cause-mortality). However, the FPC scores for the 5^th^ SBP component which discriminated higher supine SBP and impaired recovery of SBP between ∼30-70 seconds after stand were significantly associated with all-cause, circulatory system and other mortalities in multivariable analyses. One reason for this may be that the FPC scores allow for information across the time-varying haemodynamic curves to be incorporated into the model which may constitute a much richer and flexible source of information than a dichotomous or scalar summary taken at individual timepoints. Such information may also allow for subclinical features of autonomic dysregulation to be detected which may otherwise be missed by focusing on simplistic summaries such as the classical measurement of OH60.

Interestingly, despite elevated HR being a well-known risk factor for both CVD and mortality, those who were otherwise healthy at baseline but developed CVD in the intervening 12 years had lower baseline HR throughout the active stand procedure including at supine rest. However, the relative difference between those who remain CVD free and those with incident CVD is most pronounced around the initial response at the heart rate peak ∼10-20 after standing and so again may be an early indicator of autonomic dysregulation in particular over activation of the sympathetic nervous system (47).

The predominant features of autonomic dysregulation found in otherwise healthy adults who go on to have incident CVD in the intervening 12 years from our training dataset are strikingly similar to the validation cohort of those who already have CVD from the validation clinical cohort of patients referred to the Falls and Syncope Unit i.e. blunted HR peak, impaired BP recovery from ∼20 seconds after stand and elevated SVI from the stand point onwards. The fact that these features are present in those with incident CVD and also present but more pronounced in those already with a diagnosis of CVD suggests that autonomic dysregulation and the haemodynamic response to a stressor such as standing may be an important early indicator of future CVD.

We have shown that our FPCA model which was trained on a relatively healthy population representative sample of Irish adults aged 50 and over can be easily exported and used in other settings. Also, the fact that the component capturing impairment of DBP recovery to standing was significantly associated with CVD in a cohort of clinical patients with many more underlying conditions than typical in the training cohort showcases the flexibility and robustness of such a modelling approach. Using CVD as an example outcome in this case, we have demonstrated how our principal component scores, trained using TILDA data, can easily be constructed for any clinical setting which performs the active stand and also used to model any other clinical outcome of interest such as cognitive impairment or future falls.

One of the main limitations of this study is that the subsample of participants who partook in the TILDA health assessment were a much healthier subsample of the larger study and therefore the effects of peripheral haemodynamic responses to orthostasis on mortality may be underestimated in some cases. However, to our knowledge no study to date has comprehensively incorporated dynamic data driven information over the entire trace of the haemodynamic response to standing using methods such as functional data analysis to investigate incident CVD nor mortality. These component scores provide a small set of simple independent continuous measurements which capture information regarding complex trends which explain the majority of the variability in the high dimensional haemodynamic profiles and we have demonstrated how they can easily be calculated automatically in a clinical setting and incorporated into prediction models for prediction of health outcomes.

## Conclusions

This study has shown that functional principal component analysis can be used to uncover dynamic features of hemodynamic responses to standing. Our models accurately discriminated 12-year incident CVD as well as 12-year all-cause and cause specific mortality in a population cohort of adults aged 50+. We have also demonstrated how these models can be incorporated into a clinical setting and used for prediction of health outcomes, identifying impaired recovery of DBP as a significant factor associated with CVD after covariate adjustment.

To our knowledge this is the first study to incorporate dynamic data driven information over the entire trace of the haemodynamic response to standing to investigate associations with prevalent and incident CVD and mortality. The fact that components which discriminated high BP and impaired recovery of BP were associated with 12-year mortality when traditional summary measures of OH and baseline BP were not; suggests that incorporating information across the entire response to standing allows for richer and more flexible associations with health outcomes to be uncovered and allows for a better understanding of the underlying physiology leading to such associations.

## Data Availability

The dataset(s) supporting the conclusions of this article are not publicly available due to data protection regulations but are accessible at TILDA on reasonable request. The procedures to gain access to TILDA data are specified at https://tilda.tcd.ie/data/accessing-data/.

